# Malaria parasite density and detailed qualitative microscopy enhances large-scale profiling of infection endemicity in Nigeria

**DOI:** 10.1101/2022.10.18.22281220

**Authors:** Wellington Oyibo, Victoria Latham, Oladosu Oladipo, Godwin Ntadom, Perpetua Uhomoibhi, Nnnena Ogbulafor, Chukwu Okoronkwo, Festus Okoh, Aminu Mahmoud, Emmanuel Shekarau, Olusola Oresanya, Yakubu Joel Cherima, Innua Jalingo, Bintu Abba, Mohammed Audu, David J. Conway

## Abstract

With global progress towards malaria reduction stalling, further analysis of the epidemiology is required, particularly in countries with the highest burden. National surveys have mostly analysed infection prevalence, while large-scale data on parasite density and different developmental forms are rarely available. In Nigeria, the country with the largest burden globally, blood slide microscopy of children up to five years of age was conducted in the 2018 National Demographic and Health Survey, and infection prevalence previously reported. In the current study, malaria parasite density measurements are reported and analysed for 7783 of the children sampled across the 36 states within the six geopolitical zones of the country, with asexual and sexual stages considered separately and comparisons of infections with different malaria parasite species being performed. Across all states of Nigeria, there was a positive correlation between mean asexual parasite density in peripheral blood of infected individuals and prevalence of infection in the community (Spearman’s rho = 0.39, P = 0.02). Asexual parasite densities were highest in the northern geopolitical zones (geometric means > 2000 μL^-1^), extending the evidence of exceptionally high infection burden in many areas. Sexual parasite prevalence in each state was highly correlated with asexual parasite prevalence (Spearman’s rho = 0.70, P < 0.001), although sexual parasite densities were low (geometric means < 100 μL^-1^ in all zones). Infants had lower parasite densities than children above one year of age, but there were no differences between male and female children. Most infections were of *P. falciparum*, which had higher asexual densities but lower sexual parasite densities than *P. malariae* or *P. ovale* mono-infections. However, mixed species infections had the highest asexual parasite densities. It is recommended that future large surveys in high burden countries measure parasite densities as well as developmental stages and species, to improve the quality of malaria epidemiology and tracking of future changes.

## Introduction

Global progress in reducing malaria has stalled over the past several years. There were an estimated 241 million malaria cases and 627,000 deaths globally in 2020, Nigeria accounting for far more than any other country ^1^. The World Health Organization (WHO) emphasises a ‘high burden to high impact’ approach is needed for malaria control, which is most pertinent to Nigeria ^1,2^, for which a significantly improved understanding of local epidemiology will be important ^3^. Among the methods by which infection endemicity can be estimated, parasite prevalence in community surveys has been most commonly used ^4^. Other potential measurements include parasite density in the blood of infected individuals, but understanding the potential value of such data to understand variation in endemicity has not been investigated on a large scale.

National surveys of infection prevalence in children under 5 years of age in high-burden endemic countries have become important components of Malaria Indicator Surveys (MIS) or National Demographic and Health Surveys (NDHS), co-ordinated internationally by the Demographic and Health Survey (DHS) Program. Such surveys are informative, and analysis of existing data has indicated that variation in prevalence among states in Nigeria is greater than the variation within some other West African countries ^5^, highlighting the importance of analysing heterogeneity of malaria within Nigeria in more depth. Previous surveys in Nigeria have consistently shown that populations in the north of the country tend to have higher infection prevalence than those in south, and there is variation among the states within each geopolitical zone of the country ^6^.

It is not yet known known how useful parasite density estimates of infected individuals may be in large scale surveys, to add to information from prevalence data. Determinants of blood stage parasite density are complex, as acquired immunity can reduce parasite densities. It may be expected that highly endemic areas, in which people naturally acquire immunity, would have many infections with low parasite density, but a broad survey of research studies generally indicates that parasite densities within infections tend to be lower in areas of low transmission ^7^. Although many infections of low density can only be measured using highly sensitive molecular detection methods, high quality slide microscopy data are informative above a detection threshold of approximately 10 parasites per microlitre of blood. Such parasite density data can add to the power of observational research studies, as demonstrated for example in the Garki Project, an important early study of malaria epidemiology in a highly endemic area of Nigeria. In that study, infants under one year of age had a lower parasite density than children under five years of age ^8^, and children living in villages with indoor residual spraying against mosquitoes had infections of lower parasite density than children living in villages with no spraying. Most infections in Garki were due to *P. falciparum*, while *P. malariae* or *P. ovale* were also common, the probability of an individual being positive with one species being higher if another species was also detected ^8^.

Whereas the Garki Project was conducted almost 50 years ago, focusing intensively on a small area of high endemicity, it is notable that it has not been followed by broader studies on parasite density within this country. The present study is the first analysis of parasite density in a large-scale study of malaria throughout Nigeria. Combining with previous variables measured in the 2018 National Demographic and Health Survey (NDHS) of Nigeria, new data on parasite density within infections are presented for 7783 children up to five years of age, together with analysis of sexual stage parasites and the density and co-occurrence of different *Plasmodium* species. The community-based sampling of these children from all states throughout the country has enabled evaluation of the utility of these additional parasitological measurements in profiling variation in malaria infection burden.

## Methods

### Study population, blood sample collection and parasitaemia quantification

Malaria is endemic throughout Nigeria, in all six geopolitical zones and 36 states, as confirmed by the 2018 National Demographic and Health Survey (NDHS) ^9^. Natural transmission by mosquito vectors occurs in diverse environments, with vegetational zones ranging broadly from south to north of the country, ranging from mangrove forests in the coastal south, to freshwater rain forests, wooded savannas, and semi-arid savannas merging into the Sahel in the extreme north. The overall annual rainfall is higher in the south than the north of the country, but in the north a single annual peak in rainfall always occurs between July and September, whereas in southern areas there may be two seasonal peaks, timing varying from one area to another and being less consistent among years.

Under the protocol for the NDHS in 2018, children aged between 6 months and 5 years had capillary blood samples collected (finger prick samples for all children except those under one year of age for which heel prick sampling was recommended). Thick and thin peripheral blood films were prepared on the same side of uniquely barcoded slides. These slides were stained with 3% Giemsa, using the WHO-recommended malaria microscopy standard operating procedure (MM-SOP-09) at decentralised staining sites, and then transferred to a single accredited diagnostic centre with expert microscopy in Lagos, at the ANDI Centre of Excellence for Malaria Diagnosis, College of Medicine, University of Lagos. After receipt, each slide was scanned into an electronic database for slide management and data entry.

Each slide was read independently by two expert microscopists examining 200 high power fields, to determine if parasites were present. When slides were read as positive, cumulative counts of numbers of asexual parasites, sexual parasites (gametocytes) and human leukocytes were performed by the same two microscopists, examining as many fields as required until a minimum number of leukocytes were counted, at least 200 or 500 depending on the number of parasites detected. When gametocytes were detected, counts of these were performed against at least 1000 leukocytes. The estimated parasite density per microlitre of blood was calculated based on individuals containing an assumed average leukocyte count of 8000 μL^-1^. The processes and results were reviewed by a slide coordinator who checked for concordance. In the case of slides that were confirmed as positive, a consensus parasite density reading was based on the mean of the two independent estimated results if the discordance was < 20%. When the discordance in parasite counts was 20% or greater, another microscopist conducted an independent read.

### Slide read data merging, checking and curating

Excel files containing the microscopical slide read data and corresponding slide barcodes were checked for completeness, and data on the demographic information for all individuals in the NDHS 2018 survey were downloaded from the Demographic and Health Survey (DHS) database, following which the laboratory file data and DHS data were merged so that demographic information could be analysed. To merge these datasets, each file had to contain a variable in common and this was the barcode for each slide. The data were explored and checked before merging, and during this process a small number of duplicate barcode records were identified in the laboratory file which were removed before merging. After merging, only records with matching in both files were analysed (Supplementary Figure S1). Asexual density categories were created and density variables only including infected children were produced. Cross-tabulations were performed on common variables found in both the laboratory and DHS file to check data consistency prior to finalising the edited merged file for analysis.

### Statistical analyses

The distributions of malaria parasite densities among different categories were statistically compared using the non-parametric Mann-Whitney test for two categories and Kruskal-Wallis test for more than two categories. If the Kruskal-Wallis test found an overall significant difference, then pairwise comparisons using the Mann-Whitney test were conducted. Estimates of average parasite densities for each subpopulation or category were calculated as geometric means.

For the majority of the analyses on parasite density, individuals negative for parasites were excluded. As only looking at the density distribution in infected individuals meant that the analysis was not affected by prevalence differences and discrete value of density data could be determined. Where analysis was based on all individuals (positive and negative) this is stated. For selected categorical analyses, asexual densities were grouped into three categories of low (<□1000 μL^-1^) medium (1000–9999 μL^-1^), and high (10,000 μL^-1^), with Pearson’s Chi squared test used to compare proportions in each category among different subpopulations. Spearman’s non-parametric rank correlation tests were used to examine correlations between continuous variables such as asexual parasite prevalence and density. Statistical analyses were performed using STATA 16.1 and R 4.1.0.

### Ethical considerations

All the samples were collected as part of the 2018 Nationwide Multiple Indicator Cluster Survey (MICS), under the Demographic and Health Surveys (DHS) Programme (https://dhsprogram.com/). Procedures for the surveys were reviewed and approved by the ICF Institutional Review Board (IRB), and by the National Health Research Ethics Committee of Nigeria (NHREC). All methods were performed in accordance with relevant guidelines and regulations. Informed consent was obtained from all subjects or their legal guardians in the case of the children sampled for this analysis. The parasitological data generated from the microscopical analyses were merged with the anonymised demographic data from the same samples as provided by the MICS database (http://mics.unicef.org/surveys), with approval from the Ethics Committee of the London School of Hygiene and Tropical Medicine.

## Results

### Study population

After merging the detailed microscopical parasitology data with the demographic variables for the samples in the 2018 NDHS survey, complete data including parasite densities, stages and species were available for a total of 7783 children under five years of age (Supplementary Figure S1). The mean age of these children was 2.4 years, 3812 (49.0%) were females, and the numbers analysed in each of the 36 states are shown in Supplementary Table S1.

### Asexual parasite density

A total of 1675 (21.5%) children were positive for asexual malaria parasites, out of all 7783 children with complete microscopy data. Among those positive, asexual parasite densities ranged from 15 to 485,609 μL^-1^, with a geometric mean of 2242 μL^-1^ (95% CI, 2032-2473). The asexual parasite density in infected individuals was first compared among the six geopolitical zones within the country (Figure 1), as it had previously been shown that malaria prevalence varied geographically, northern zones having generally higher prevalence than in the south ^6,9^. This revealed higher geometric mean asexual parasite densities in infected individuals in the zones in the North than those in the South of the country (Figure 1). The highest density was in the North West zone (geometric mean 2650 μL^-1^, 95% CI 2229-3151) and the lowest in the South West zone (geometric mean 1663 μL^-1^, 95% CI 1296-2133). There was highly significant variation among geopolitical zones overall (Kruskal-Wallis test, P < 0.001), and pairwise comparisons showed that the North West and North Central zones each had significantly higher parasite densities within infections than each of the Southern zones (Mann-Whitney tests, P < 0.01).

**Figure 1.**
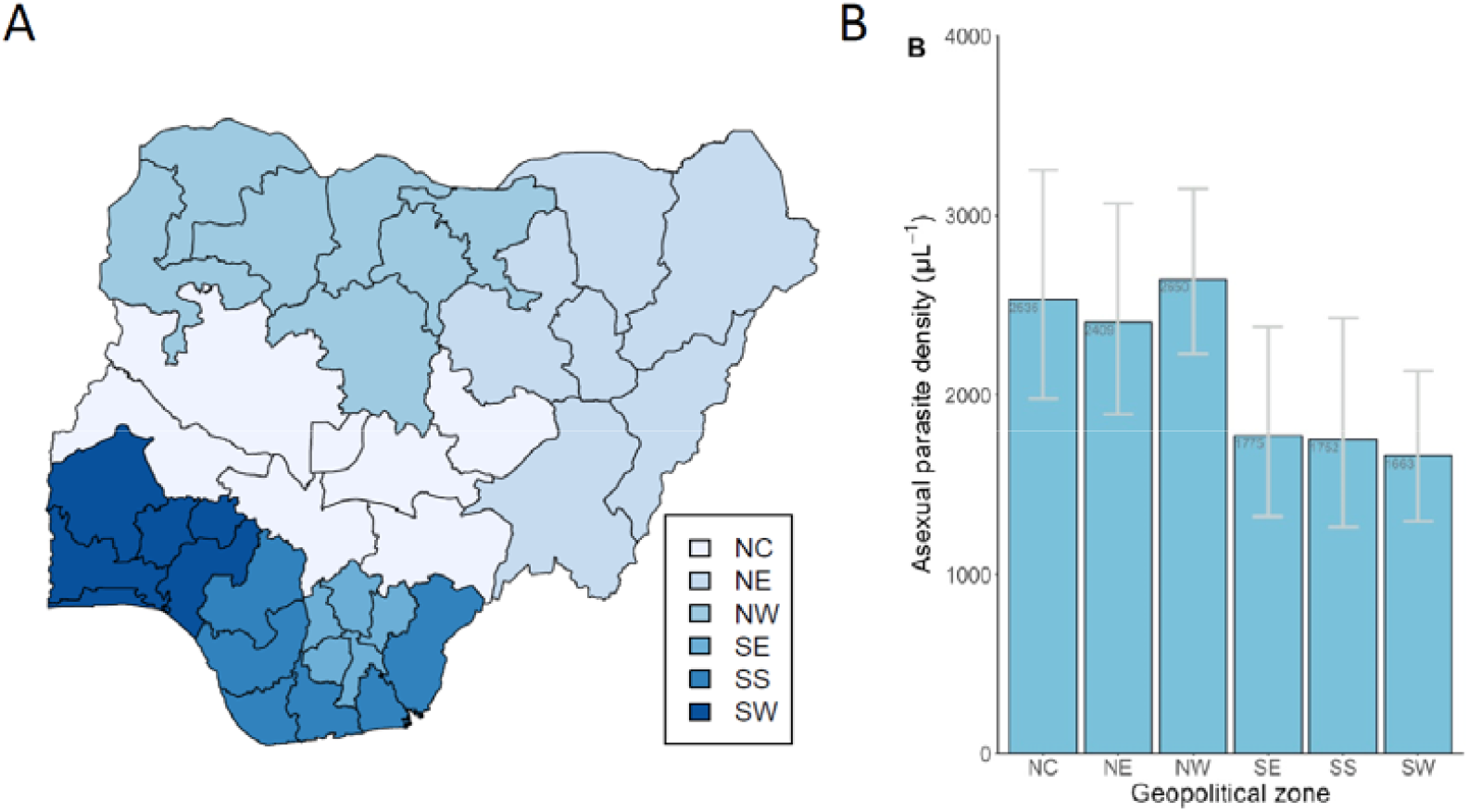
Malaria parasite densities within infections in children up to 5 years of age vary among the major geopolitical regions of Nigeria. **A**. Map of Nigeria, showing the major six geopolitical zones of the country, and the individual states (map produced in R using the naijR package). **B**. Geometric mean parasite densities (with 95% confidence intervals) among slide-positive children in the six different major geopolitical zones (NC, North Central; NE, North East, NW, North West; SE, South East; SS, South South; SW, South West). There is significant overall heterogeneity (Kruskal-Wallis test, P < 0.001), and each zone in the north of the country has higher density infections than each zone in the south of the country (Mann-Whitney pairwise tests, P < 0.001).

Although there were higher asexual parasite densities within infections in the northern zones of the country, there was also significant variation in the mean parasite densities among different states within each geopolitical zone (Figure 2A and Supplementary Table S1). Therefore, analysis was conducted to investigate the correlation between mean parasite density within infections for each state and the prevalence of infection in each state (Figure 2B and Supplementary Table S1). This revealed a significant positive correlation (Spearman’s r = 0.39, P = 0.02), indicating that states with a higher infection prevalence tended to have higher asexual parasite density within infections (Figure 2C).

**Figure 2.**
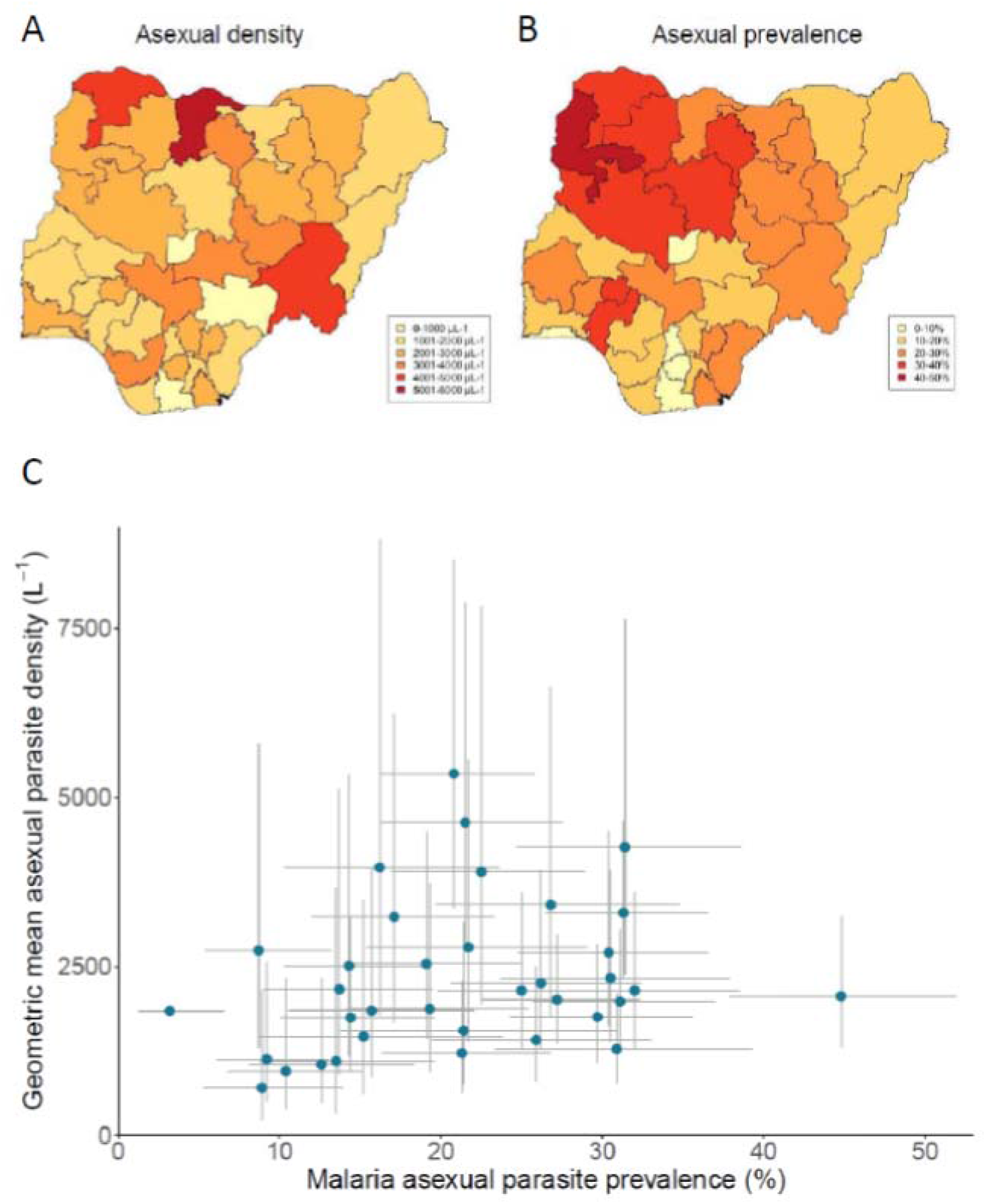
Geographical variation in mean malaria parasite density in infected children under five years of age in Nigeria. **A**. Variation in geometric mean parasite density among all 36 states, densities shaded by value categories as indicated. The names of individual states are shown in Figure 1, and numerical values (with 95% CIs of estimates) are tabulated in Supplementary Table S1. **B**. Prevalence of malaria parasite infection in each of the states, shaded by value categories as indicated, as previously analysed ^6^. **C**. Scatterplot showing a positive significant correlation between asexual parasite prevalence and density within infections in each state in Nigeria. Lines indicate the 95% CI for the density and prevalence for each state (a point on the far left corresponds to Lagos which does not show 95% CI for density as there were too few samples positive for accurate estimation). All values are shown in Supplementary Table S1.

Overall, asexual parasite densities of infected individuals did not differ between males (geometric mean 2187 μL^-1^, 95% CI 1909-2505) and females (geometric mean 2304 μL^-1^, 95% CI 1998-2657, P > 0.5) (Table 1). The prevalence of asexual parasite infection varied by age, ranging from 13.3% in those under one year of age to 28.1% in those aged five years (Table 1 and Supplementary Figure S2), and among those who were infected there was also variation in asexual parasite density with age (Kruskal-Wallis test, p=0.04). Pairwise comparisons between age categories showed that this difference is primarily due to those under one year of age having lower densities (geometric mean 1414 μL^-1^, 95% CI 985-2030) compared to the other children (Table 1 and Supplementary Figure S3).

**Table 1.** Asexual malaria parasite prevalence and density according to sex, age and geopolitical zone in 7783 children up to five years of age with complete microscopy data for all variables.

|  | <u>Prevalence</u> |  |  | <u>Density in infected individuals</u> |  |  |
| --- | --- | --- | --- | --- | --- | --- |
| | Asexual parasite prevalence (%) <sup>a</sup> | 95% CI (%) | P value <sup>b</sup> | Asexual parasite density Geometric mean ($\mu\text{L}^{-1}$) | 95% CI ($\mu\text{L}^{-1}$) | P value <sup>c</sup> |
| **Sex** |  |  |  |  |  |  |
| Male | 22.2 (882/3971) | 20.9-23.5 | 0.1 | 2187 | 1909-2505 | 0.5 |
| Female | 20.8 (793/3812) | 19.5-22.1 |  | 2304 | 1998-2657 |  |
| **Age (years)** |  |  |  |  |  |  |
| < 1 | 13.3 (100/754) | 10.9-15.9 | <0.001 | 1414 | 985-2030 | 0.04 |
| 1 | 17.4 (285/1637) | 15.6-19.3 |  | 1928 | 1494-2487 |  |
| 2 | 17.7 (290/1638) | 15.9-19.6 |  | 2750 | 2161-3499 |  |
| 3 | 25.2 (441/1752) | 23.2-27.3 |  | 2383 | 1986-2858 |  |
| 4 | 27.9 (461/1653) | 25.7-30.1 |  | 2197 | 1816-2659 |  |
| 5 | 28.1 (98/349) | 23.4-33.1 |  | 2538 | 1698-3795 |  |
| **Geopolitical zone** |  |  |  |  |  |  |
| North-Central | 20.7 (283/1368) | 18.6-22.9 | <0.001 | 2535 | 1977-3251 | <0.001 |
| North-East | 20.1 (280/1391) | 18.1-22.3 |  | 2409 | 1893-3065 |  |
| North-West | 30.8 (554/1797) | 28.7-33.0 |  | 2650 | 2229-3151 |  |
| South-East | 15.4 (186/1207) | 13.4-17.6 |  | 1775 | 1322-2382 |  |
| South-South | 15.3 (130/852) | 12.9-17.9 |  | 1752 | 1264-2429 |  |
| South-West | 20.7 (242/1168) | 18.4-23.2 |  | 1663 | 1296-2133 |  |

<sup>a</sup> Note that the prevalence numbers have slightly different denominators from those in the original published report on the DHS prevalence data<sup>9</sup>, as only slides with complete data for all variables including parasite density are analysed here
<sup>b</sup> Pearson's chi squared test
<sup>c</sup> Mann-Whitney or Kruskal-Wallis tests

### Sexual stage parasite prevalence and density

Sexual stage parasites (gametocytes) were detected in 604 (7.8%) of the 7783 children having complete microscopy data. The observed gametocyte prevalence varied significantly by age, being lowest in infants under one year of age (4.9%, 95% CI 3.5-6.7%) (Table 2). The gametocyte prevalence also varied among geopolitical zones within the country, being higher in the northern zones than in the south (Table 2). Across all states, there was a strong positive significant correlation between gametocyte prevalence and asexual parasite prevalence, as expected (Spearman’s r = 0.70, P < 0.001)(Figure 3).

**Table 2.** Sexual parasite prevalence and density according to sex, age and geopolitical zone in 7783 children up to five years of age with complete microscopy data for all variables.

|  | <u>Prevalence</u> |  |  | <u>Density in infected individuals</u> |  |  |
| --- | --- | --- | --- | --- | --- | --- |
| | Sexual parasite prevalence (%) | 95% CI (%) | P value <sup>a</sup> | Sexual parasite density Geometric mean ($\mu\text{L}^{-1}$) | 95% CI ($\mu\text{L}^{-1}$) | P value <sup>b</sup> |
| **Sex** |  |  |  |  |  |  |
| Male | 8.2 (324/3971) | 7.3-9.1 | 0.20 | 85 | 74-98 | 0.33 |
| Female | 7.3 (280/3812) | 6.5-8.2 |  | 77 | 66-90 |  |
| **Age (years)** |  |  |  |  |  |  |
| < 1 | 4.9 (37/717) | 3.5-6.7 | <0.001 | 71 | 47-107 | 0.23 |
| 1 | 6.3 (103/1534) | 5.2-7.6 |  | 89 | 69-114 |  |
| 2 | 7.6 (124/1514) | 6.3-9.0 |  | 91 | 70-118 |  |
| 3 | 8.8 (154/1598) | 7.5-10.2 |  | 65 | 54-79 |  |
| 4 | 9.7 (160/1493) | 8.3-11.2 |  | 90 | 74-110 |  |
| 5 | 7.4 (26/323) | 4.9-10.7 |  | 80 | 45-141 |  |
| **Geopolitical zone** |  |  |  |  |  |  |
| North Central | 8.8 (120/1248) | 7.3-10.4 | <0.001 | 94 | 75-119 | 0.63 |
| North East | 7.0 (97/1294) | 5.7-8.4 |  | 89 | 66-121 |  |
| North West | 12.1 (217/1579) | 10.6-13.7 |  | 77 | 65-91 |  |
| South East | 4.3 (52/1155) | 3.2-5.6 |  | 69 | 48-98 |  |
| South South | 4.3 (37/815) | 3.1-5.9 |  | 75 | 48-117 |  |
| South West | 7.8 (81/1087) | 5.5-8.5 |  | 79 | 60-104 |  |

<sup>a</sup> Pearson's chi squared test
<sup>b</sup> Mann-Whitney or Kruskal-Wallis tests

**Figure 3.**
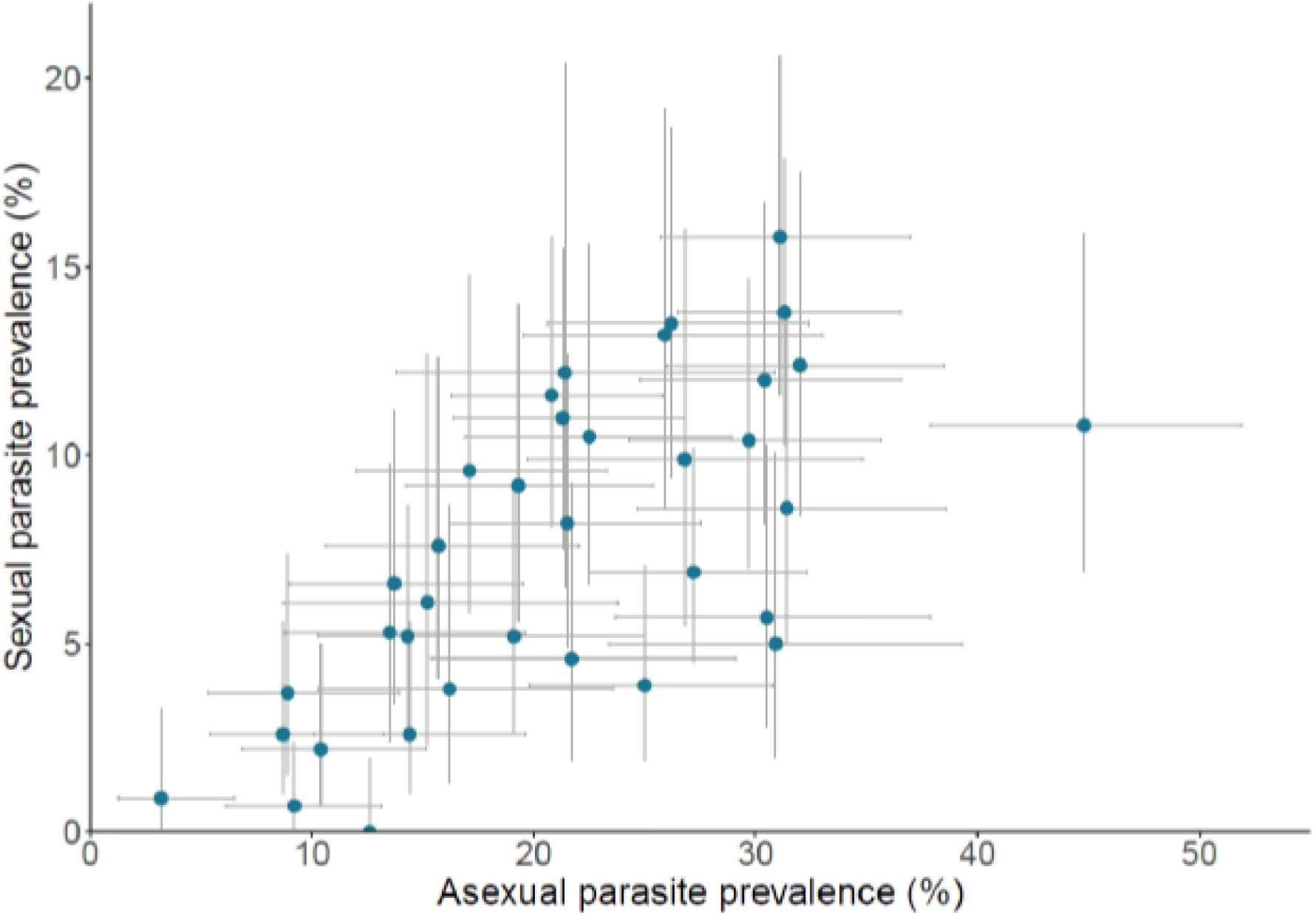
Significant positive correlation between asexual and sexual malaria parasite prevalence in each state (Spearman’s rho = 0.70, P < 0.001). 95% CIs are shown for all estimates, and all values are shown in Supplementary Table S1.

Among those individuals that were detected as gametocyte-positive, the geometric mean gametocyte density was 81 μL^-1^ (95% CI 73-90). There were no significant differences in gametocyte densities across the different age categories, or the different geopolitical zones in Nigeria, among those that were positive (Table 2). Although gametocyte prevalence is an epidemiologically informative measure, the estimated gametocyte density in positive individuals may not offer a very useful additional measurement. In most infections with sexual parasites detected, their numbers are not very far above the microscopical detection threshold, thereby giving the density estimates relatively low precision, in contrast to the asexual parasites which have much higher numbers counted in most infections.

### Different malaria parasite species

Most (87.6%) of the slide-positive malaria parasite infections had *P. falciparum* detected alone, while 3.9% of infections had *P. malariae* alone, 1.0% had *P. ovale* alone. The remainder were mixed species infections, with 6.0% of all infections having both *P. falciparum* and *P. malariae* detected, while 1.3% had both *P. falciparum* and *P. ovale*, and no infection was seen to contain both *P. malariae* and *P. ovale* (Table 3). Although much less common than *P. falciparum*, the other species *P. malariae* and *P. ovale* were detected in the samples from all zones and most states throughout Nigeria (Supplementary Table S2).

**Table 3.** Prevalence and density of different Plasmodium species infections in children up to 5 years of age in Nigeria.

| *Plasmodium* species detected | Asexual parasite prevalence (%; N = 7783) | Asexual density <sup>a</sup>, parasites $\mu\text{L}^{-1}$ (95% CI) | Sexual parasite prevalence (%; N = 7783) | Sexual density <sup>a</sup>, parasites $\mu\text{L}^{-1}$ (95% CI) |
| --- | --- | --- | --- | --- |
| *P. falciparum* alone | 18.6<br>(1450) | 2304<br>(2066-2570) | 6.1<br>(472) | 67<br>(59-75) |
| *P. malariae* alone | 0.9<br>(71) | 982<br>(718-1343) | 0.6<br>(45) | 185<br>(142-242) |
| *P. ovale* alone | 0.2<br>(19) | 622<br>(322-1202) | 0.1<br>(11) | 173<br>(71-421) |
| *P. falciparum* & *P. malariae* mixed infection | 1.4<br>(109) | 3021<br>(2298-3971) | 0.8<br>(61) | 153<br>(121-194) |
| *P. falciparum* & *P. malariae* mixed infection | 0.3<br>(24) | 3984<br>(2282-6955) | 0.2<br>(15) | 141<br>(74-270) |
|  |  | P < 0.001 <sup>b</sup> |  | P < 0.001 <sup>b</sup> |

<sup>a</sup> Geometric means (with 95% confidence intervals) of parasite densities tabulated.
<sup>b</sup> Kruskal-Wallis tests for comparisons of parasite densities across categories.
Slide read data on all individuals are given in Supplementary Table S3.

There were highly significant differences in asexual parasite densities among different *Plasmodium* species infections (p<0.001, Figure 4 and Table 3). Single species infections with *P. falciparum* alone had much higher asexual parasite densities (geometric mean 2304 μL^-1^) than infections with *P. malariae* alone (geometric mean 982 μL^-1^) or *P. ovale* alone (geometric mean 622 μL^-1^). However, co-infections with *P. falciparum* and either of the other species had higher asexual parasite densities (geometric mean 3021 μL^-1^ for *P. falciparum* and *P. malariae* co-infections, and 3984 μL^-1^ for *P. falciparum* and *P. ovale* co-infections) (Figure 4A, Table 3). In contrast to the differences in densities for asexual parasites, in *P. malariae* and *P. ovale* single species infections there were higher sexual stage parasite densities than for *P. falciparum* infections (Figure 4B, Table 3).

**Figure 4.**
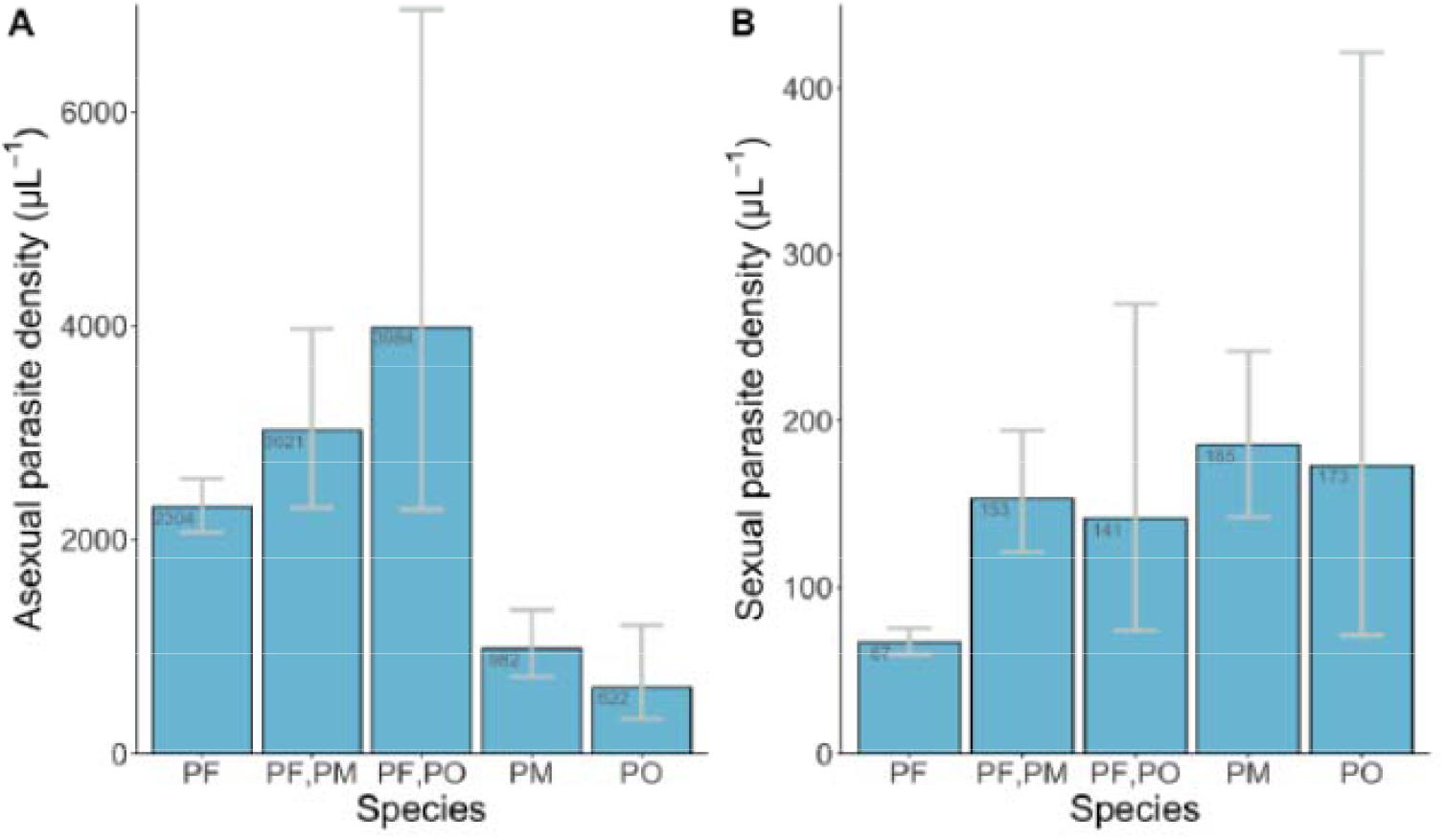
Geometric means (with 95% CIs) of asexual and sexual parasite densities for different *Plasmodium* species and co-infections in Nigeria (PF, *P. falciparum*; PM, *P. malariae*; PO, *P. ovale*; no slide was positive for *P. vivax*). Sample sizes (numbers of infections with asexual parasites counted) for the asexual density measurements were 1450, 109, 24, 71 and 19 (for PF, PF/PM, PF/PO, PM and PO respectively). Sample sizes (numbers of infections with sexual parasites counted) for the sexual density measurements was 472, 61, 15, 45 and 11 (for PF, PF/PM, PF/PO, PM and 19 respectively).

## Discussion

Parasite prevalence has been most frequently used to estimate population variation in malaria infection burden, but parasite density and other microscopical measurements can add information that increases epidemiological discrimination. Parasite density measurements were utilised for understanding variation in community infection burden in the Garki Project in northern Nigeria almost 50 years ago ^8^, but the current study is the first description of parasite densities nationwide, enabling a powerful large-scale analysis across diverse areas. This could encourage more targeted interventions aimed at reducing the parasite density in particular areas which comprise major reservoirs for malaria transmission.

The results indicate that parasite density yields additional information on variation in parasite infection burden. Prevalence data and asexual density within infections correlate at the state level, with high prevalence areas tending to have higher parasite densities than low prevalence areas, although there is a range of densities in each setting covering several orders of magnitude. It is important to note that data in this study are from children up to five years of age, and that there will be other determinants of parasite densities in older children or adults who have acquired varying levels of immunity in different areas. Different hypothesises have been suggested to explain varying parasite densities in areas with different levels of malaria transmission, with potentially complex interactions between causes ^7,10,11^. It is known that malaria parasite densities vary greatly over time in individual infections, as shown from cohort studies of natural infections in endemic populations ^12^, and artificially induced infections ^13^, but analysing distributions of parasite densities in a population sample is informative for comparative studies.

Consistent with results from community-based surveys elsewhere in Africa, this large study shows lower levels of parasite density in infections in infants under one year of age than in other children up to five years of age ^8,14^. This may be partly due to maternal antibodies, and foetal haemoglobin which may restrict the parasite densities ^15^, although these will have a greater effect in those under six months of age who were not sampled in this study. Moreover, infants tend to have less exposure due to their lower mobility ^16^, being wrapped and remaining close to their mothers, and having a small surface area for mosquito biting ^17^. As the age distribution of children sampled was similar in all states ^9^, the slightly lower parasite density generally seen in infants does not systematically affect any analyses comparing across the different states.

There are limits to estimating sexual stage parasite densities using microscopy, given their normally low density. Previous studies using molecular techniques have indicated that most individuals with asexual parasites also contain sexual parasites, but they are often not detected by microscopy due to their low density ^18^, and when detected the small numbers lead to considerable stochastic sampling variation in density estimations. It has been shown that having a higher density of sexual stages in the blood is correlated with increased transmission, although low densities are also able to infect mosquitoes ^19^. In *P. falciparum*, developing sexual stages sequester in the bone marrow, so only very early or late-stage sexual stages are present in the bloodstream ^20,21^, further limiting the density estimations.

Most malaria infections of the young children in this study contained *P. falciparum*, and the small minority with either *P. malariae* or *P. ovale* alone had generally lower parasite densities in the blood. If a more sensitive detection method were used it is likely that a higher prevalence of these non-falciparum species would be detected. Although these species are less virulent than *P. falciparum*, they ought not to be neglected in ongoing malaria control, as in some other areas where *P. falciparum* has substantially reduced there has been an apparent increase in the proportions of one or more of these species ^22,23^. It is possible that some antimalarials are more effective in clearing *P. falciparum* than other species ^24,25^, and *P. ovale* parasites are better able to persist during treatment due to their dormant liver stage that enables relapse, with chronic infections potentially causing anaemia in asymptomatic children ^25^. *P. falciparum* is also more likely to cause symptomatic infections that are treated, compared to the other species which have much lower density in the blood. It has previously been suggested that *P. falciparum* has a suppressive effect on *P. malariae* ^26^, although in the current study *P. falciparum* and *P. malariae* coinfections had higher parasite densities than infections with either species alone.

Previous studies on sexual stage parasites in co-infections of different species have shown variable results, either indicating that co-infecting species limit *P. falciparum* sexual stage production or that they enhance production ^27,28^. It has been suggested that the sequence of the co-infections alters the dynamics, with sexual stage production is only enhanced if *P. malariae* infection occurs before or at the same time as *P. falciparum* ^28^. The potential effect of co-infections in modifying risk of clinical disease also varies among studies ^29-31^, and correlations between co-infections and parasite densities have varied between areas of a single country ^32^.

Parasite density data adds information to that provided by detected prevalence in young children, but it needs to be considered how such information may be best obtained and utilised. One issue is whether to use microscopy or molecular methods for parasite detection and density estimation. The usefulness of slide microscopy may be relatively high in areas of high transmission but of limited power in areas of low transmission. For example, in community surveys in West African countries with lower malaria endemicity than in Nigeria, PCR assays reliably detect several-fold more infections than microscopy ^33^, and these are now used more commonly for malaria epidemiological research in these populations. In Nigeria, molecular methods would also give more sensitive estimation of the non-falciparum malaria infection burden, as illustrated by recent local studies in the south of the country ^34,35^.

Based on the analyses here, it would be useful for future nationwide Demographic and Health Surveys to generate data on parasite density in positive individuals, to go beyond the basic measure of infection prevalence. For example, such information could help assess the effect of seasonal malaria chemoprevention interventions in northern states of Nigeria, as parasite density reduction could be incorporated with other indices including prevalence and numbers of reported clinical cases. However, data should be comparable among different surveys, as well as within a single nationwide survey, which requires standardisation of protocols. An example of a methodological issue for attention is the potential for batch effects, where drying, staining or humidity sometimes affect the slide preparations ^36^. Comparisons are more straightforward if samples are processed in the same laboratory using the same protocol ^37^, as in the present study. By designing other surveys knowing they will be compared, it is possible to minimise systematic biases in broader comparisons, whether utilising microscopy or molecular techniques. A commitment to such data generation is needed, as more focus on parasite density would enhance surveillance in high burden populations where most future malaria control efforts are required.

## Data Availability

All data generated or analysed during this study are included in this manuscript and its supplementary information files.

## Author contributions

All authors contributed to conceptualisation and conduct of the study; W.O., O.Ol., G.N., P.U., N.O., C.O., F.O., A.M., E.S., O.Or., Y.J.C., I.J., B.A. and M.A. were responsible for sample collection, survey materials and data; W.O. managed the generation of laboratory slide microscopy data; V.L., W.O., D.J.C. performed data analyses; W.O., V.L., D.J.C. wrote the manuscript; all authors advised on drafts and approved the final version of the manuscript.

## Data availability

All data generated or analysed during this study are included in this published article and its supplementary information files.

## References

1 WHO. World Malaria Report 2021. (Geneva, 2021).

2 WHO. High burden to high impact: a targeted malaria response. https://www.who.int/malaria/publications/atoz/high-impact-response/en/. (World Health Organization, Geneva, 2019).

3 Rosenthal, P. J., John, C. C. & Rabinovich, N. R. Malaria: How are we doing and how can we do better? Am J Trop Med Hyg 100, 239–241, doi:10.4269/ajtmh.18-0997 (2019).

4 Hay, S. I., Smith, D. L. & Snow, R. W. Measuring malaria endemicity from intense to interrupted transmission. Lancet Infect Dis 8, 369–378, doi:10.1016/S1473-3099(08)70069-0 (2008).

5 Abeles, J. & Conway, D. J. The Gini coefficient as a useful measure of malaria inequality among populations. Malar J 19, 444, doi:10.1186/s12936-020-03489-x (2020).

6 Oyibo, W. et al. Geographical and temporal variation in reduction of malaria infection among children under 5 years of age throughout Nigeria. BMJ Glob Health 6, doi:10.1136/bmjgh-2020-004250 (2021).

7 Okell, L. C. et al. Factors determining the occurrence of submicroscopic malaria infections and their relevance for control. Nat Commun 3, 1237, doi:10.1038/ncomms2241 (2012).

8 Molineaux, L. & Gramiccia, G. The Garki project: research on the epidemiology and control of malaria in the Sudan Savanna of West Africa. (World Health Organization, 1980).

9 National Population Commission (NPC) [Nigeria] and ICF International. Nigeria Demographic and Health Survey 2018. (NPC and ICF, Abuja, Nigeria, and Rockville, Maryland, USA, 2019).

10 Bousema, T., Okell, L., Felger, I. & Drakeley, C. Asymptomatic malaria infections: detectability, transmissibility and public health relevance. Nat Rev Microbiol 12, 833–840, doi:10.1038/nrmicro3364 (2014).

11 Slater, H. C. et al. The temporal dynamics and infectiousness of subpatent Plasmodium falciparum infections in relation to parasite density. Nat Commun 10, 1433, doi:10.1038/s41467-019-09441-1 (2019).

12 Delley, V. et al. What does a single determination of malaria parasite density mean? A longitudinal survey in Mali. Trop Med Int Health 5, 404–412, doi:10.1046/j.1365-3156.2000.00566.x (2000).

13 O’Meara, W. P., Collins, W. E. & McKenzie, F. E. Parasite prevalence: a static measure of dynamic infections. Am J Trop Med Hyg 77, 246–249 (2007).

14 D’Alessandro, U. et al. Malaria in infants aged less than six months - is it an area of unmet medical need? Malar J 11, 400, doi:10.1186/1475-2875-11-400 (2012).

15 Billig, E. M., McQueen, P. G. & McKenzie, F. E. Foetal haemoglobin and the dynamics of paediatric malaria. Malar J 11, 396, doi:10.1186/1475-2875-11-396 (2012).

16 McElroy, P. D. et al. Predicting outcome in malaria: correlation between rate of exposure to infected mosquitoes and level of Plasmodium falciparum parasitemia. Am J Trop Med Hyg 51, 523–532 (1994).

17 Goncalves, B. P. et al. Examining the human infectious reservoir for Plasmodium falciparum malaria in areas of differing transmission intensity. Nat Commun 8, 1133, doi:10.1038/s41467-017-01270-4 (2017).

18 Bousema, T. & Drakeley, C. Epidemiology and infectivity of Plasmodium falciparum and Plasmodium vivax gametocytes in relation to malaria control and elimination. Clin Microbiol Rev 24, 377–410, doi:10.1128/CMR.00051-10 (2011).

19 Churcher, T. S. et al. Predicting mosquito infection from Plasmodium falciparum gametocyte density and estimating the reservoir of infection. Elife 2, e00626, doi:10.7554/eLife.00626 (2013).

20 Aguilar, R. et al. Molecular evidence for the localization of Plasmodium falciparum immature gametocytes in bone marrow. Blood 123, 959–966, doi:10.1182/blood-2013-08-520767 (2014).

21 Joice, R. et al. Plasmodium falciparum transmission stages accumulate in the human bone marrow. Sci Transl Med 6, 244re245, doi:10.1126/scitranslmed.3008882 (2014).

22 Yman, V. et al. Persistent transmission of Plasmodium malariae and Plasmodium ovale species in an area of declining Plasmodium falciparum transmission in eastern Tanzania. PLoS Negl Trop Dis 13, e0007414, doi:10.1371/journal.pntd.0007414 (2019).

23 Akala, H. M. et al. Plasmodium interspecies interactions during a period of increasing prevalence of Plasmodium ovale in symptomatic individuals seeking treatment: an observational study. Lancet Microbe 2, e141–e150, doi:10.1016/S2666-5247(21)00009-4 (2021).

24 Gneme, A. et al. Plasmodium species occurrence, temporal distribution and interaction in a child-aged population in rural Burkina Faso. Malar J 12, 67, doi:10.1186/1475-2875-12-67 (2013).

25 Sutherland, C. J. Persistent parasitism: The adaptive biology of malariae and ovale malaria. Trends Parasitol 32, 808–819, doi:10.1016/j.pt.2016.07.001 (2016).

26 Ritchie, T. L. Interactions between malaria parasites infecting the same vertebrate host. Parasitology 96, 607–639 (1988).

27 Bousema, J. T. et al. Increased Plasmodium falciparum gametocyte production in mixed infections with P. malariae. Am J Trop Med Hyg 78, 442–448 (2008).

28 McKenzie, F. E., Jeffery, G. M. & Collins, W. E. Plasmodium malariae infection boosts Plasmodium falciparum gametocyte production. Am J Trop Med Hyg 67, 411–414, doi:10.4269/ajtmh.2002.67.411 (2002).

29 Black, J., Hommel, M., Snounou, G. & Pinder, M. Mixed infections with Plasmodium falciparum and P. malariae and fever in malaria. Lancet 343, 1095 (1994).

30 May, J. et al. Impact of subpatent multi-species and multi-clonal plasmodial infections on anaemia in children from Nigeria. Trans R Soc Trop Med Hyg 94, 399–403, doi:10.1016/s0035-9203(00)90119-6 (2000).

31 Smith, T. et al. Prospective risk of morbidity in relation to malaria infection in an area of high endemicity of multiple species of Plasmodium. Am J Trop Med Hyg 64, 262–267, doi:10.4269/ajtmh.2001.64.262 (2001).

32 Bruce, M. C. et al. Effect of transmission setting and mixed species infections on clinical measures of malaria in Malawi. PLoS One 3, e2775, doi:10.1371/journal.pone.0002775 (2008).

33 Satoguina, J. et al. Comparison of surveillance methods applied to a situation of low malaria prevalence at rural sites in The Gambia and Guinea Bissau. Malar J 8, 274 (2009).

34 Oriero, E. C. et al. Seroprevalence and parasite rates of Plasmodium malariae in a high malaria transmission setting of southern Nigeria. Am J Trop Med Hyg 103, 2208–2216, doi:10.4269/ajtmh.20-0593 (2020).

35 Abdulraheem, M. A. et al. High prevalence of Plasmodium malariae and Plasmodium ovale in co-infections with Plasmodium falciparum in asymptomatic malaria parasite carriers in southwestern Nigeria. Int J Parasitol 52, 23–33, doi:10.1016/j.ijpara.2021.06.003 (2022).

36 Bejon, P. et al. Thick blood film examination for Plasmodium falciparum malaria has reduced sensitivity and underestimates parasite density. Malar J 5, 104, doi:10.1186/1475-2875-5-104 (2006).

37 O’Meara, W. P. et al. Sources of variability in determining malaria parasite density by microscopy. Am J Trop Med Hyg 73, 593–598 (2005).

