## Supplementary Figure for "Malaria parasite density and detailed qualitative microscopy enhances large-scale profiling of infection endemicity in Nigeria"

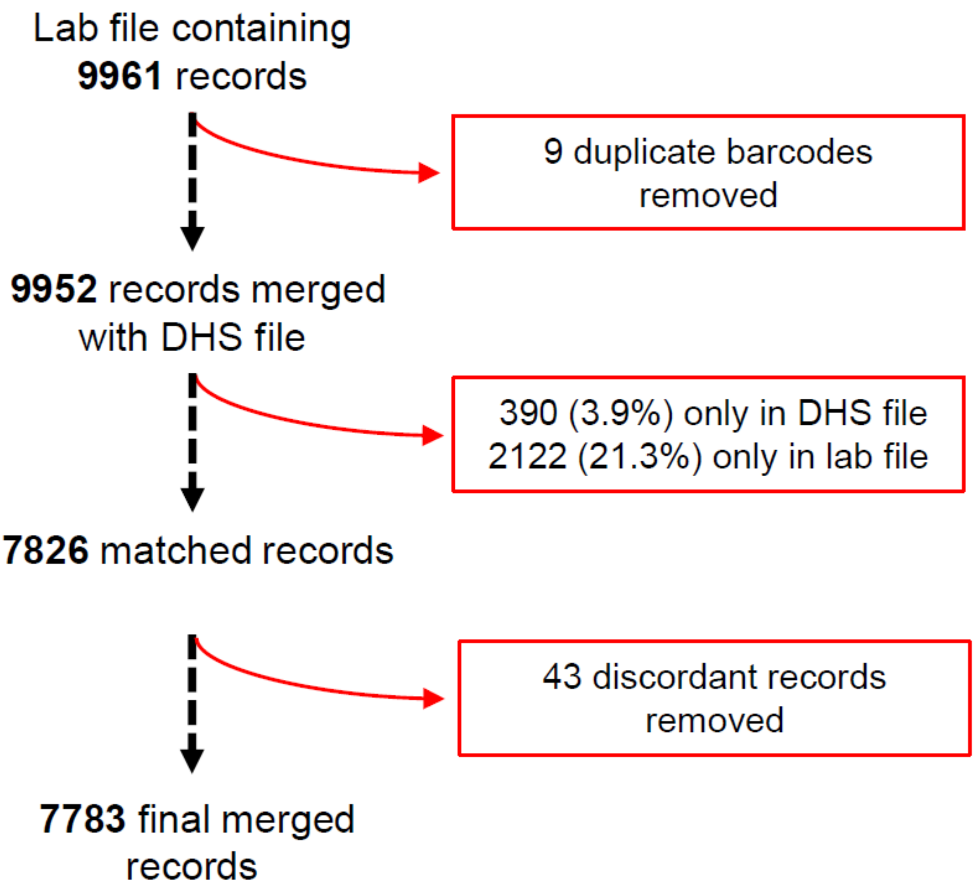


**Supplementary Figure S1.** Flow-diagram of sample filtering and database merging for the laboratory microscopy data on children up to 5 years of age sampled in the 2018 NDHS survey.

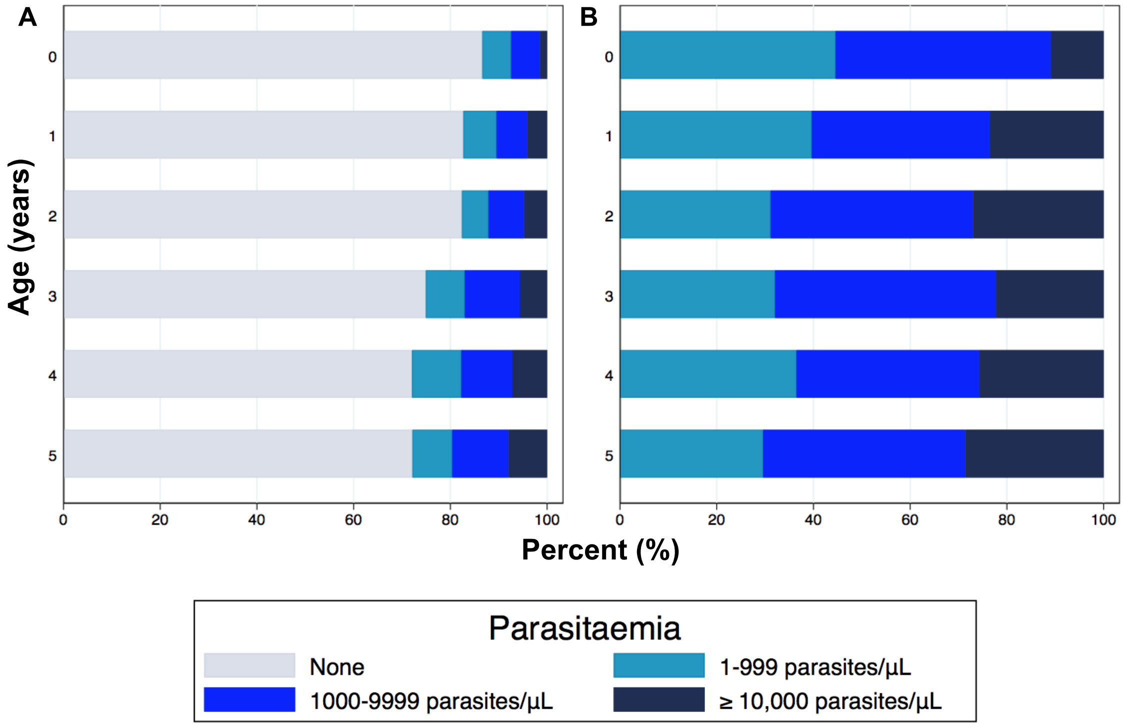


**Supplementary Figure S2.** Asexual parasite density categories of malaria parasite infections in different age groups. **A.** All individuals included in the analysis (including slide negative with zero counts as well as slide positive). **B.** Analysis of slide-positive individuals.

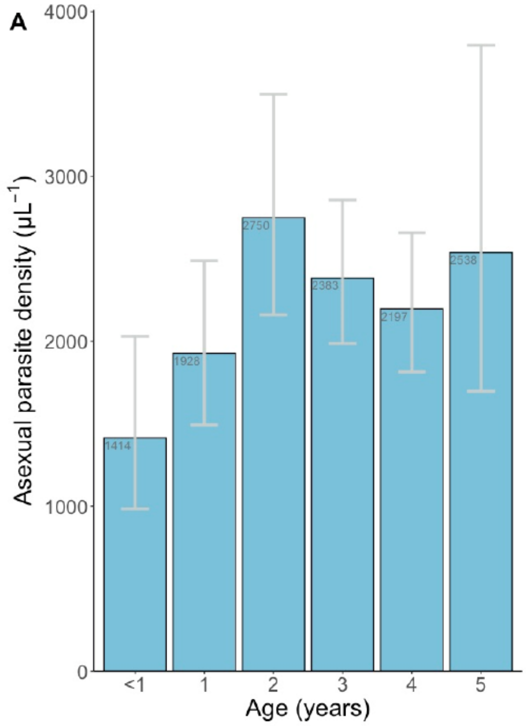


**Supplementary Figure S3.** Geometric mean parasite densities (with 95% CIs) among slide-positive individuals of different ages. Overall there was significant variation by age (Kruskal-Wallis test, p=0.04), which was mostly attributable to lower densities in infants under one year of age.
